# Shape variation and sexual dimorphism of the adult human mandible evaluated by geometric morphometrics

**DOI:** 10.1101/2023.11.18.23298726

**Authors:** Aspasia Chalazoniti, Wanda Lattanzi, Demetrios J. Halazonetis

**Affiliations:** Department of Prosthodontics, School of Dentistry, National and Kapodistrian University of Athens, Greece; Department of Life Science and Public Health, Università Cattolica del Sacro Cuore, Rome, Italy; Fondazione Policlinico Universitario A. Gemelli IRCCS, Rome, Italy; Department of Orthodontics, School of Dentistry, National and Kapodistrian University of Athens, Greece

**Keywords:** Mandible, shape, sexual dimorphism, geometric morphometrics, atlas

## Abstract

Mandibular shape variability and effects of age and sex were explored in an adult human sample using dense landmarking and geometric morphometrics. We segmented 50 male and 50 female mandibular surfaces from CBCT images (age range: 18.9 to 73.7 years). Nine fixed landmarks and 496 sliding semilandmarks were digitized on the mandibular surface, and then slid by minimizing bending energy against the average shape. Principal component analysis extracted the main patterns of shape variation. Sexes were compared with permutation tests and allometry was assessed by regressing on the log of the centroid size. Almost 49 percent of shape variation was described by the first three principal components. Shape variation was related to width, height and length proportions, variation of the angle between ramus and corpus, height of the coronoid process and inclination of the symphysis. Significant sexual dimorphism was detected, both in size and shape. Males were larger than females, had a higher ramus, more pronounced gonial angle, larger inter-gonial width, and more distinct antegonial notch. Accuracy of sexing based on the first two principal components in form space was 91 percent. The degree of edentulism was weakly related to mandibular shape. Age effects were not significant.

## Introduction

Understanding human mandibular form and its variations, beyond its biological significance, is clinically useful, particularly in cases of osseous defects, arising from trauma, pathology, congenital abnormalities, or, more frequently, loss of teeth and consequent alveolar bone resorption. Reconstruction of such defects can be guided by symmetry, if the contralateral side is present, or based on an atlas of normal variation that is adapted to the remaining parts [1, 2, 3]. This approach is commonly followed in the anthropology literature, where severely damaged specimens are reconstructed by thin plate spline (TPS) warping of a reference template [4]. A dense landmark configuration and comprehensive coverage of the specimen are obviously essential for detailed and accurate results.

Although mandibular shape has been studied extensively, both in 2-dimensional and 3-dimensional (3D) form, the morphology of the bone does not provide a large number of clearly identifiable landmarks for comprehensive coverage of its surface, potentially leading to loss of important phenotypic information. However, even if achievable, dense landmarking would not be sufficient unless complemented with reasonable confidence of landmark homology (correspondence) across specimens.

We recognize two main approaches for establishing correspondences and landmarking of 3D surface meshes; both use a reference template (atlas) with the landmarks of interest already identified on it. The first approach performs a rigid alignment of the template mesh to match the target, followed by deformable registration to refine the match, then transfers the landmarks from the atlas to the target mesh. Examples are ALPACA [5, 6] and MeshMonk [7, 8], which mainly differ in their non-rigid registration algorithm. Variants of this approach abolish landmarks altogether and achieve correspondences between the vertices of the meshes directly [3, 9].

The second approach needs digitization of a (relatively small) subset of the landmarks on both meshes, then performs a TPS warping of the template, driven by this subset of landmarks, and transfers the remaining points to the target mesh, usually by projection on the mesh surface. Thus, this is a TPS warping of point configurations whereas the first approach is essentially a mesh-to-mesh registration. The second method belongs to the toolbox of geometric morphometric (GM) methods [4, 10, 11] and is usually followed by sliding of semilandmarks to enhance geometric correspondence [12, 13] and mapping of the specimens in a shape space via Generalized Procrustes alignment (GPA) [14] and Principal Component Analysis (PCA). The shape space is the final goal of all methods, as it represents a statistical shape model [2, 3] which describes population variability and can be used both as a reference for testing novel shapes and as a generative source for creating plausible anatomy.

Studies of the mandible with dense landmarking are scarce. Most studies limit the landmarks (either static or sliding) on the anterior and posterior ramus ridges, the mandibular notch, the inferior outline, and the symphysis outline on the midsagittal plane, in addition to ubiquitous conventional landmarks, such as Gonion, Gnathion, Coronoid and the condylar poles [15, 16, 17, 18, 19, 20, 21]. The total number of landmarks ranges from below 20 (e.g. [18]) to above 100 (e.g. 113 [17] or 301 [20]); however, seldom is the mandibular surface between ridges landmarked [6, 22, 23].

The aim of this study was to create a 3D atlas of human mandibular shape variability and explore sex and age effects in an adult sample using a dense landmark configuration, uniformly covering the mandibular surface.

## Materials and Methods

### Ethical approval

All methods were carried out in accordance with relevant guidelines and regulations. The Scientific Committee of the Dental Association of Attica, Greece, and the Research Ethics Committee of the School of Dentistry, National and Kapodistrian University of Athens, gave ethical approval for this work (protocol 109777). Informed consent was obtained from all subjects involved in the study.

### Sample

The sample consisted of cone-beam computed tomography (CBCT) images, in the form of DICOM files, from the archives of a dental imaging center and the Department of Oral Diagnosis and Radiology of the School of Dentistry, the National and Kapodistrian University of Athens. The data had been obtained previously for diagnostic purposes, unrelated to this study and all patients had signed consent forms for use of their images for research purposes. Voxel size ranged between 0.2 and 0.4 mm. The images were anonymized, retaining only age and sex data. For this study, we excluded patients who presented with gross anatomical deformities (e.g. hemifacial microsomia, missing condyle or ramus) but included patients with normal morphological variations, missing or extracted teeth, and alveolar bone resorption due to tooth loss.

We accrued patients until the sample included 50 adult males and 50 adult females of comparable age distribution (Table 1, Figure 1). This size is considered sufficient for obtaining reliable data using geometric morphometric tools [24, 25]. Seventy-six patients had at least one tooth missing, excluding the third molars, with an average number of 2.9 missing teeth among them (Table 1).

**Figure 1.**
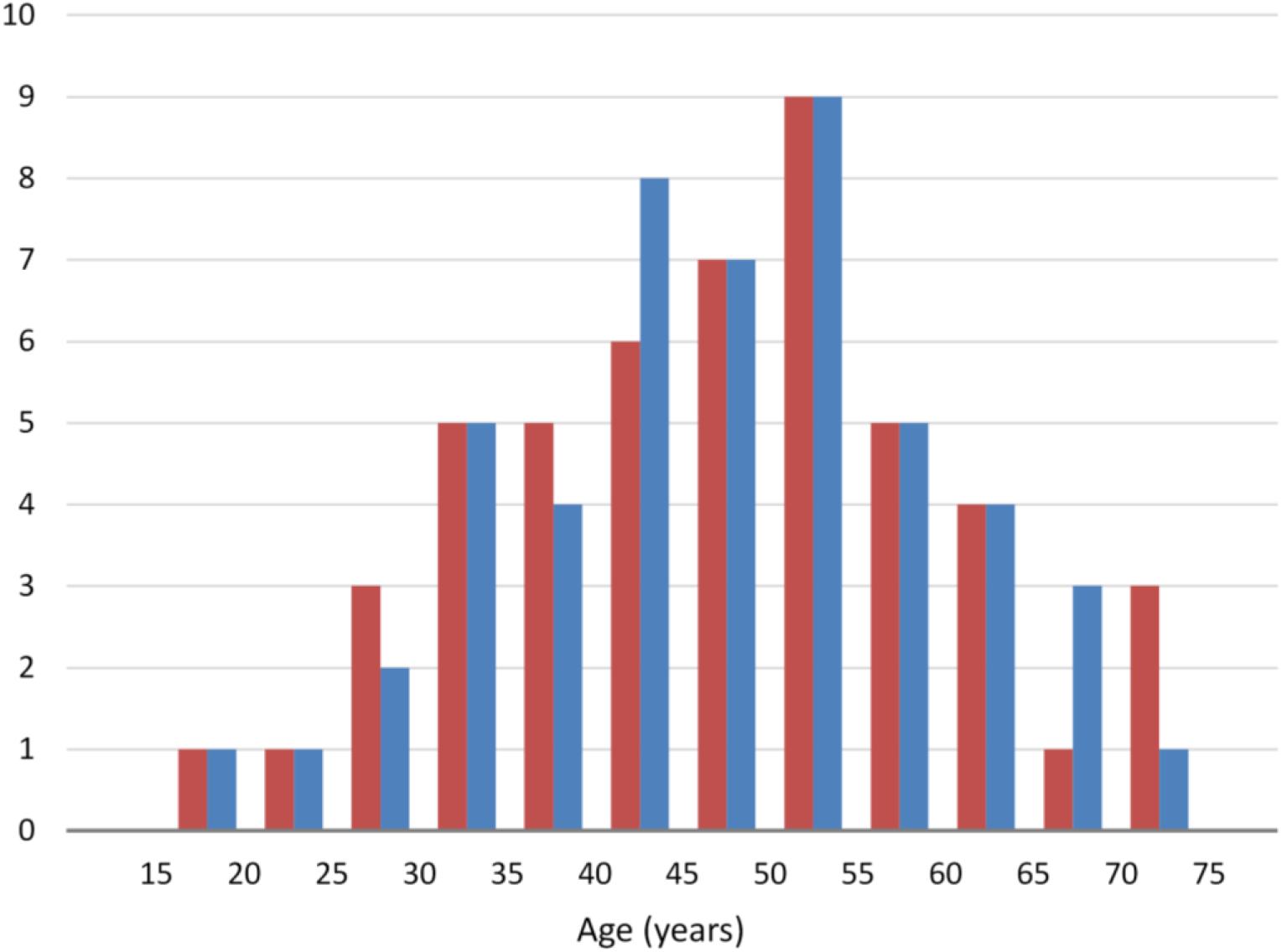
Histogram of age (years) for the female (red) and male (blue) groups.

**Table 1.** Demographics of the sample.

|  | Females | Males |
| --- | --- | --- |
| Count | 50 | 50 |
| Age (years) |  |  |
| Average (Standard Deviation) | 46.9 (13.2) | 47.3 (12.1) |
| Median | 48.6 | 48.2 |
| Range | 18.9 to 73.7 | 19.1 to 71.0 |
| Missing teeth |  |  |
| Total teeth missing | 104 | 115 |
| Maximum per patient | 9 | 11 |
| Number of patients with missing teeth | 37 | 39 |

### Preprocessing

All data processing and analysis was performed using the Viewbox 4 software (dHAL Software, Kifissia, Greece). The DICOM files were loaded and the volume subsampled by 50% for faster processing, if voxel size was 0.2 mm or smaller. The histogram of the voxel values was cropped by setting the top 0.2% of the voxels to the 99.8% value, to enhance contrast. The bone threshold was computed using Otsu multilevel histogram thresholding [26] with three levels (air, soft tissues and bone). Segmentation was based on the computed bone threshold and manually refined at each axial slice using the software’s paintbrush, by the same observer (AC). Due to frequent artifacts from metallic restorations and prostheses, teeth could not always be segmented with confidence, and tooth surfaces should be considered unreliable. A triangular mesh surface was subsequently constructed from the segmentation using a variant of the marching cubes algorithm [27]. The internal bone structure was deleted and holes were closed, obtaining a water-tight mesh of the mandibular surface, including the dentition. Finally, a remeshing filter was applied, to increase triangle regularity and remove degenerate triangles [28]. The final meshes averaged approximately 130.000 vertices and 260.000 triangles. The average edge length of the triangles was 0.44 mm. All surfaces were uploaded and are available from the Zenodo repository (https://doi.org/10.5281/zenodo.7882821).

### Landmarking

Digitization of the meshes was based on a template of curves and landmarks, with an even distribution over the entire mandibular surface, except for the dentition (Figure 2, Supplementary Table 1). The template was based on a previously used template of 415 landmarks [22, 23], refined and augmented. We used a simplified symmetric mandibular mesh as the base for placing the landmarks. The mesh was modified by expanding it outwards by 1.5 mm, thus making it thicker overall, but most importantly in the ramus region (Supplementary Figure 1). This avoided a common problem of the landmarks projecting on the wrong side of a thin ramus, and made ‘inflating’ the landmarks along the normals to the surface unnecessary [29]. The curves were cubic splines, adjusted to the mandibular surface by control points, and used for sliding the curve semilandmarks. There were 9 fixed landmarks, 84 curve semilandmarks and 426 surface semilandmarks, free to slide over the mesh surface, for a total of 519 points.

**Figure 2.**
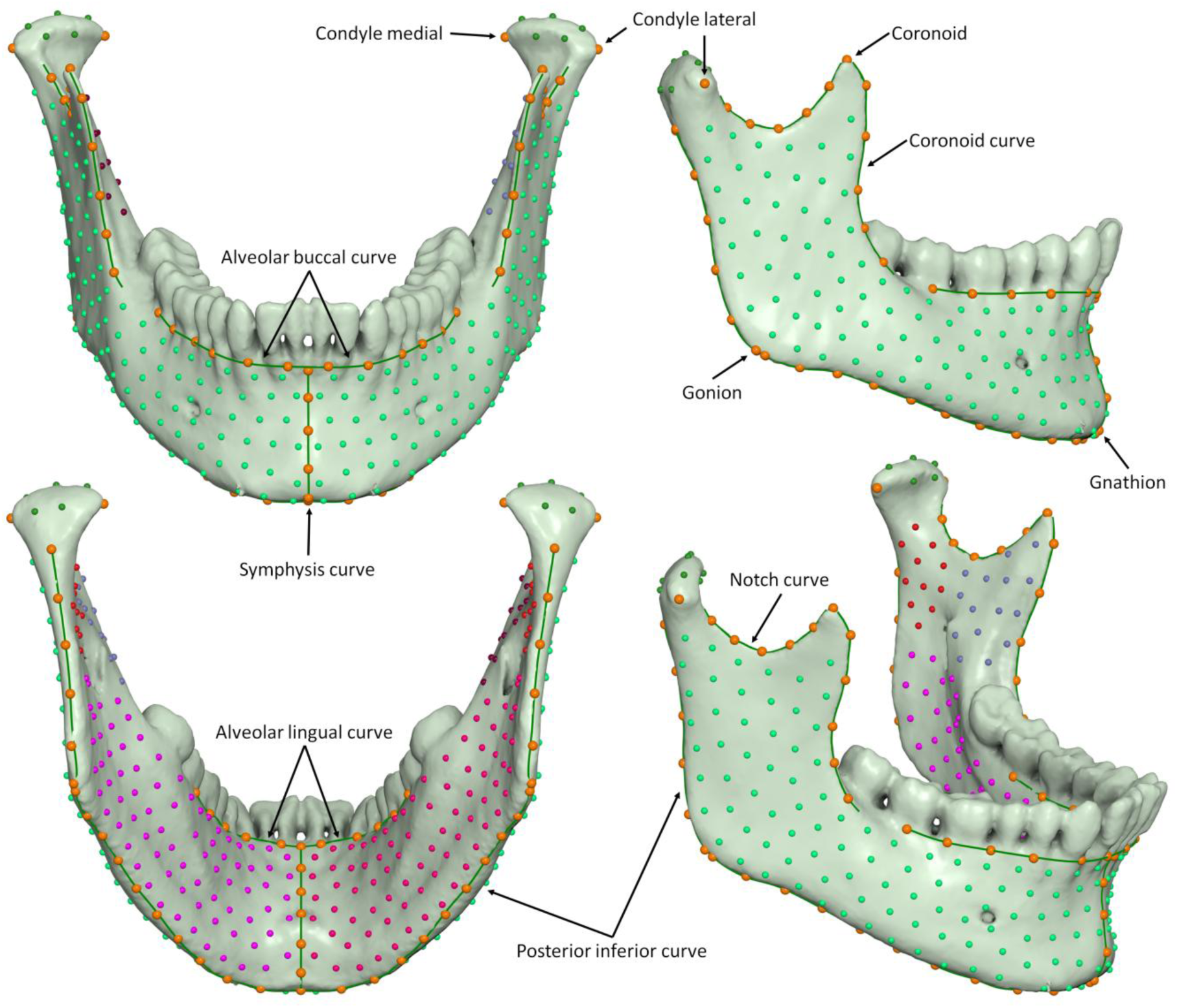
Curves and landmarks shown on one of the meshes of the sample.

Landmarks Condyle lateral, Condyle medial, Gonion and Gnathion were automatically located by the software as extrema of the mesh surface along pre-specified directions (Supplementary Table 1). The curves Notch, Coronoid, and Posterior-inferior were also located automatically based on mesh curvature data and extremal directions. The Alveolar buccal and Alveolar lingual curves were digitized manually to follow the alveolar bone crest. All other landmarks were initially placed by a Thin Plate Spline (TPS) warping of the template to the previously digitized points and curves, and then projected on the closest point of the mesh surface. Digitizations were inspected and corrected manually. All digitizations were performed by the same investigator (AC).

### Geometric Morphometrics

Shape analysis was based on the traditional toolbox of 3D geometric morphometric (GM) methods. After each mesh was digitized, the semilandmarks were allowed to slide against the sample average shape, to minimize bending energy [12, 13, 30], then all landmarks were re-projected on their corresponding curves or on the mesh surface. Sliding and projecting was repeated five times, each time reducing the sliding step in a linear fashion, to avoid oscillations in landmark position. The average shape was then re-computed and used as the reference configuration for a subsequent iteration of sliding and projecting. All configurations were then aligned using the Generalized Procrustes alignment (GPA) method [11], centroid size was computed, and a Principal Component Analysis (PCA) was run to extract the most significant shape variation patterns as Principal Components (PC). PCA was performed both in shape space and form space, which includes the logarithm of the centroid size - ln(CS) - as an extra variable [31].

Additionally, we computed the volume enclosed by the mesh surface, and calculated the mesh normalized centroid size as the square root of the average sum of the squared distances of the mesh vertices to their centroid.

In order to investigate potential sexual dimorphism in the area of the gonial angle and ramus, we ran a regional GPA and PCA analysis, confined to the landmarks of the ramus, on the right side. This included a total of 123 landmarks (Supplementary Figure 2).

### Statistical analysis

Digitization error was tested by repeating the digitization of 20 randomly selected meshes by a second investigator (DJH) and comparing the Procrustes distances between repeats to the extent of the sample in shape space.

Shape patterns were visualized by TPS warping the average shape along the PC directions over the range of ±3 standard deviations (SD). The distribution of the sample was inspected in shape space by a 3-dimensional plot of the relevant PCs. Comparison between the sexes was based on the Procrustes distance between the group means, using permutation tests.

Sexual shape dimorphism was visualized by TPS warping of the average shape along the trajectory connecting the average male and female shapes, also exaggerated three times for clarity. Discriminant analysis was used to compute the percentage of correctly classified subjects by sex, based on shape and size. Static allometry was tested by regression of the ln(CS) on the shape PCs. Sexual dimorphism in mandibular size was also tested by comparing the mandibular volumes and mesh centroid sizes.

Geometric morphometric tests and visualizations were performed by Viewbox; other statistical tests were performed using StatsDirect 1.8.10 (StatsDirect Ltd, Wirral, UK) and PAST 4.09 [32].

## Results

### Digitization error

The average Procrustes distance between the 20 repeated digitizations was 2.9% of the extent of the sample in shape space, as determined by double the distance of the farthest specimen from the centre of the sample (diameter of hypersphere enclosing the sample). The repeated digitizations virtually coincided with the originals in the PC1-PC2 plot (Supplementary Figure 3), showing that digitization error was not a consideration in the analysis.

### Shape space analysis

The plot of the sample in shape space is shown in Figure 4. The first 4 PCs described 57% of the shape variance; the first 12 PCs described 80% of the variance (Supplementary Table 2). Shape variation, as described by the 3 first PCs, is shown in Figure 5. PC1 described variability in the width of the mandible, the height of the ramus and the alveolar process. PC2 described variability in the angle between the ramus and the corpus, whereas PC3 described variability in the height of the coronoid process, the inclination of the symphysis and the prominence of the mandibular angle.

**Figure 3.**
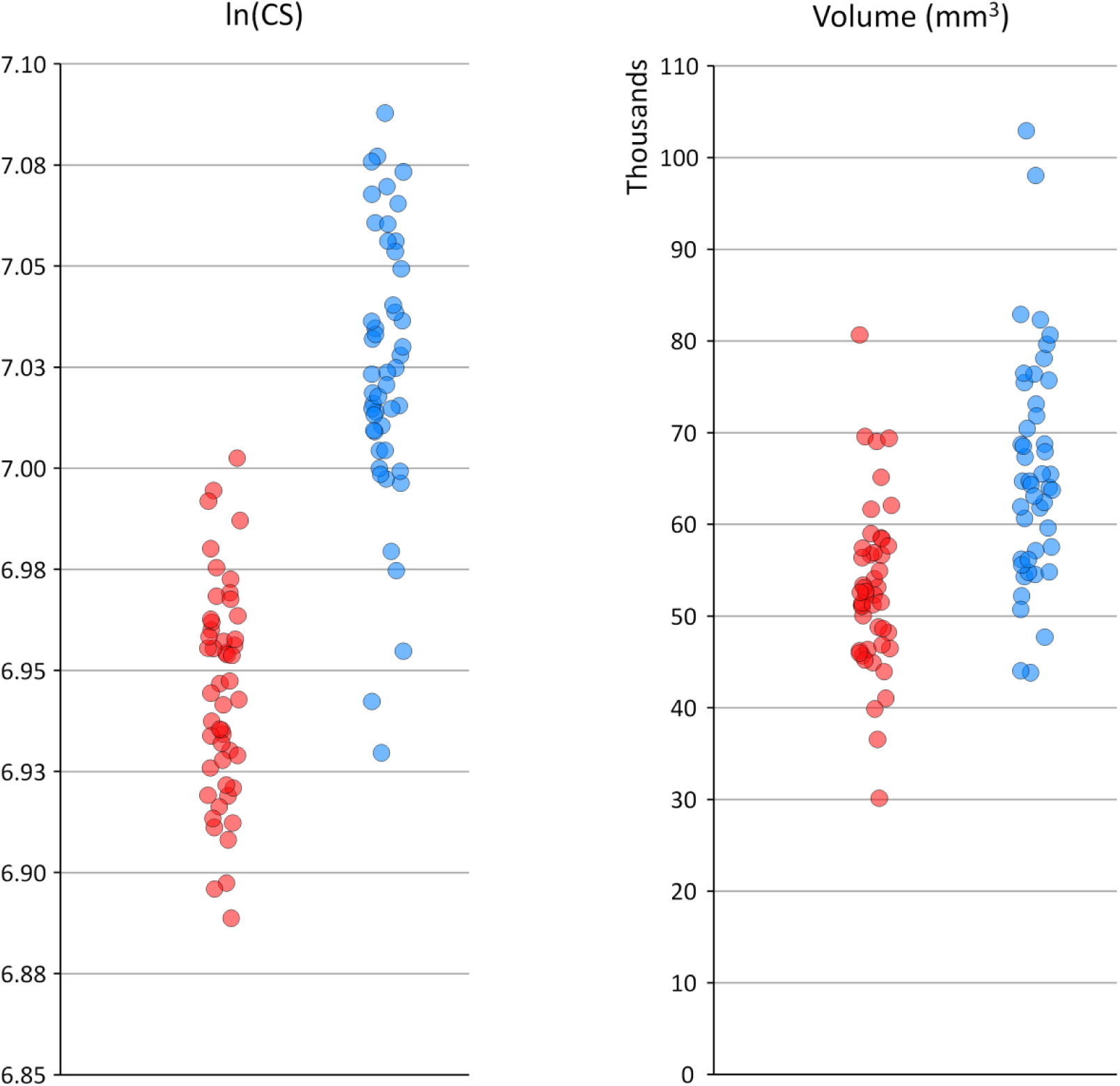
Jitter strip plots of ln(CS) and mesh volume for the female (red) and male (blue) groups.

**Figure 4.**
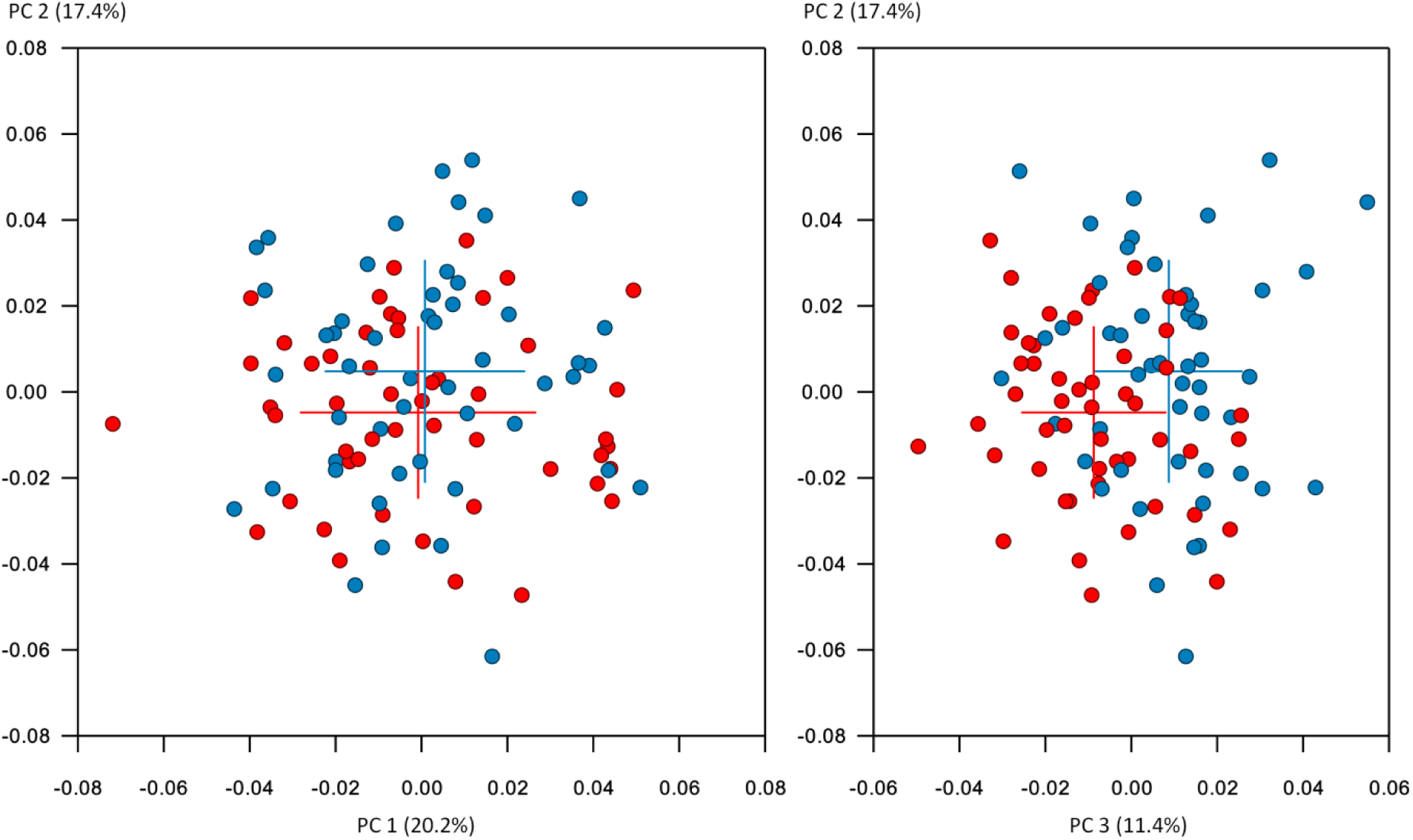
The sample plotted in shape space. Red: females, blue: males. Crosses show average of each group and standard deviations.

**Figure 5.**
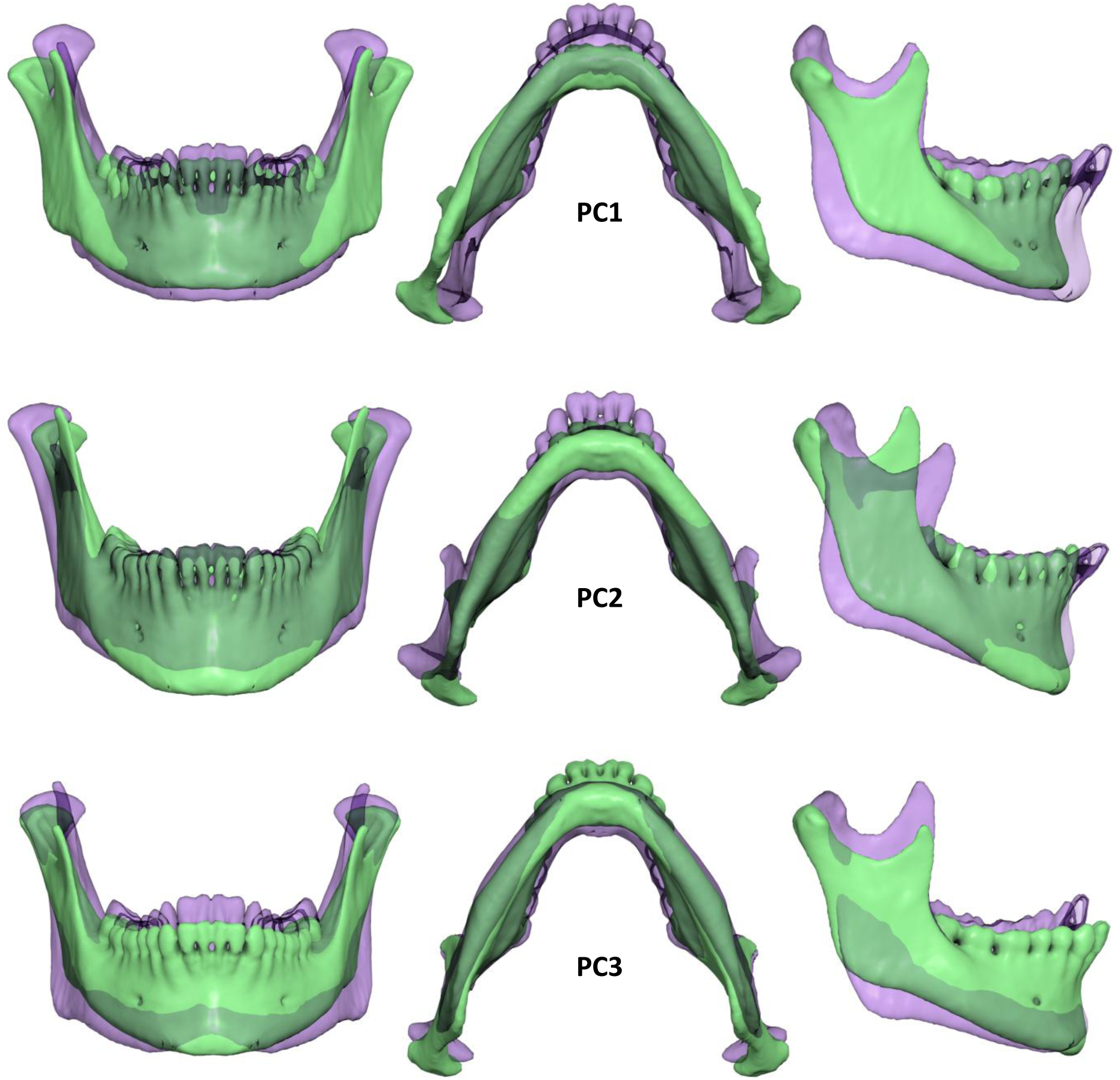
Shape variations described by the first 3 PCs. Average shape warped along each PC at -3 and +3 standard deviations (purple and green).

### Sexual dimorphism

The male and female groups differed significantly in shape (100,000 permutations on Procrustes distance, P < 0.0002), mainly along PC2 and PC3. Figure 6 shows the intergroup shape difference of the average male and female shapes, exaggerated 3 times along the male-female vector, keeping the same centroid size. Males showed a wider mandible at the gonial angles, but narrower at the condyles and coronoids, a higher ramus with higher condylar and coronoid processes, more pronounced antegonial notch, and a more posteriorly inclined symphysis with prominence at menton. In contrast, females had a larger mandibular angle, a wider ramus and a more gracile body. These are relative differences, with size adjusted to be equal between the two groups.

**Figure 6.**
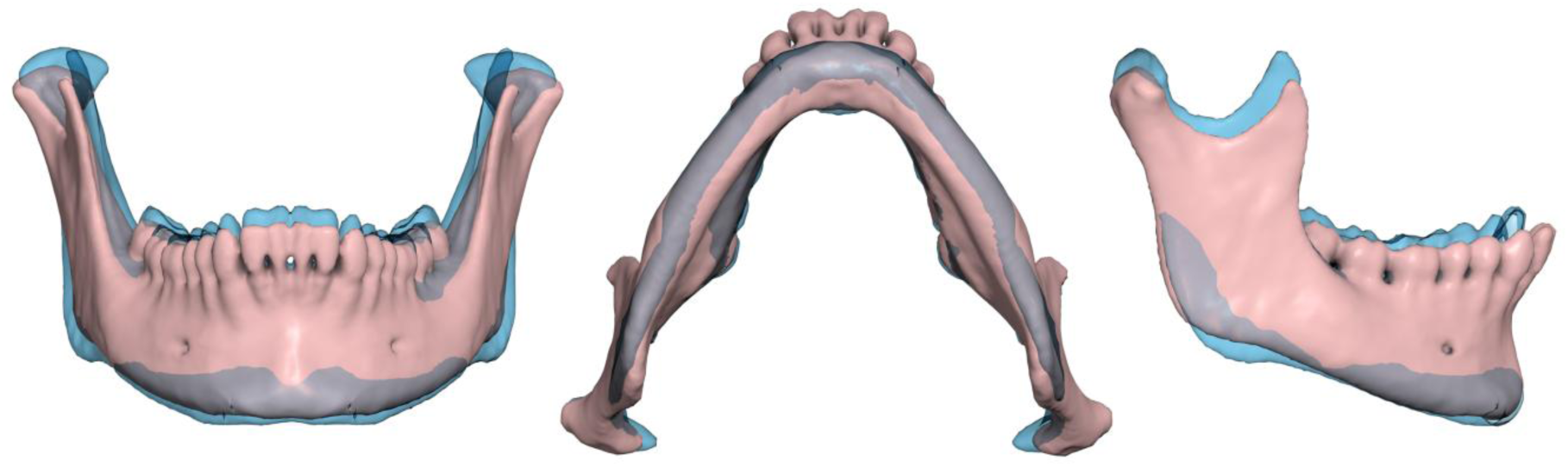
Superimposition of the average male and female shapes exaggerated 3 times along the male-female shape vector to enhance differences. Both shapes are scaled to the same centroid size. Male: blue, female: red.

There was a clear sexual dimorphism in mandibular size, expressed as the mesh centroid size (males larger by 8.9%), the mandibular volume (25%), or the landmark centroid size (8.3%) (Table 2, Figure 3). The plot of the sample in form space showed very little overlap (Supplementary Figure 4).

**Table 2.**
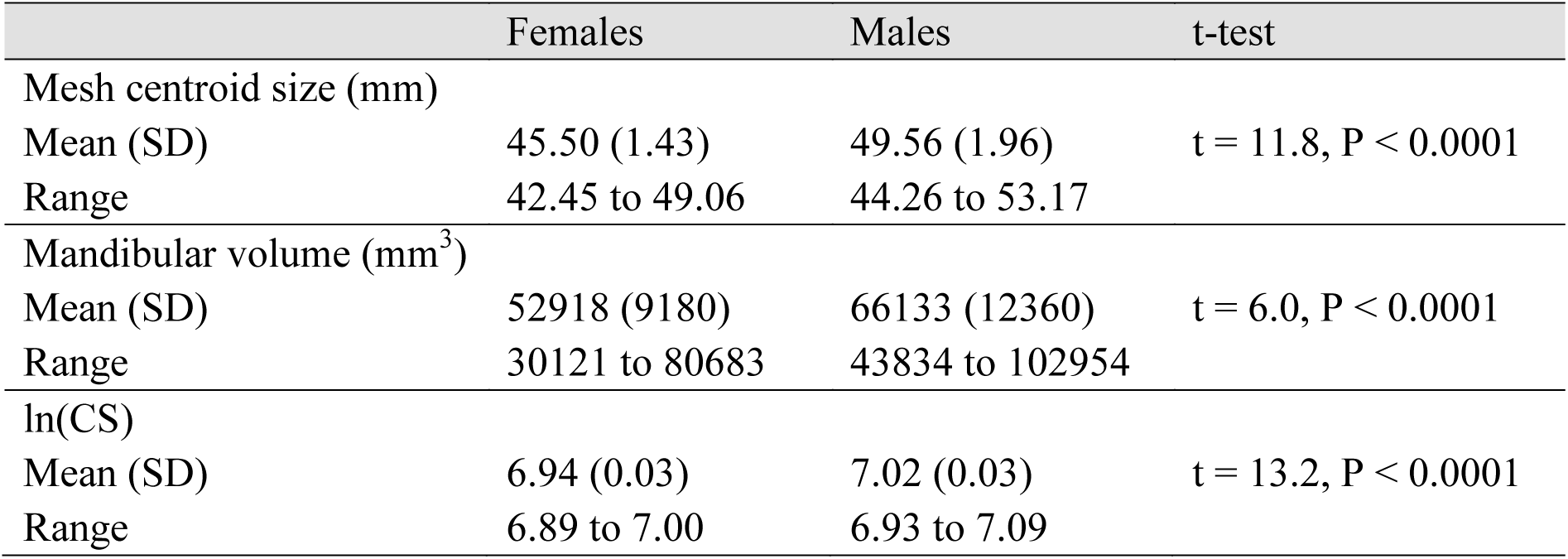
Size variables. Student’s t-test.

|  | Females | Males | t-test |
| --- | --- | --- | --- |
| Mesh centroid size (mm) |  |  |  |
| Mean (SD) | 45.50 (1.43) | 49.56 (1.96) | $t = 11.8, P < 0.0001$ |
| Range | 42.45 to 49.06 | 44.26 to 53.17 |  |
| Mandibular volume (mm<sup>3</sup>) |  |  |  |
| Mean (SD) | 52918 (9180) | 66133 (12360) | $t = 6.0, P < 0.0001$ |
| Range | 30121 to 80683 | 43834 to 102954 |  |
| ln(CS) |  |  |  |
| Mean (SD) | 6.94 (0.03) | 7.02 (0.03) | $t = 13.2, P < 0.0001$ |
| Range | 6.89 to 7.00 | 6.93 to 7.09 |  |

The correlation between volume and centroid size was weak to moderate, even within each group (Pearson correlation coefficient, females: r^2^ = 0.16, P = 0.0038, males: r^2^ = 0.21, P = 0.0007). This is partly explained by the substantial effect on volume that tooth loss and ensuing alveolar bone resorption in the vertical dimension have, without significantly affecting the average distance of the landmarks from the centroid, and hence the centroid size.

Discriminant analysis using the first 4 principal components in form space (where PC1 is heavily weighted on centroid size) showed a 93% correct sex classification (using a leave-one-out cross-validation procedure). This reduced to 91% when including only PC1 and PC2.

### Static allometry

Regression of ln(CS) on the shape variables of the pooled sample showed a significant but low correlation, less than 5% of the shape variance explained by size (10,000 permutations, predicted variance = 4.44%, P < 0.0001). However, this was lost when regressing each group separately (Table 3). In contrast, mandibular volume was related to the shape variables in both groups separately, and in the pooled sample, explaining almost 13% of shape variance in the female group. The number of missing teeth was also related to shape, but only in the male group and the pooled sample (Table 3).

**Table 3.**
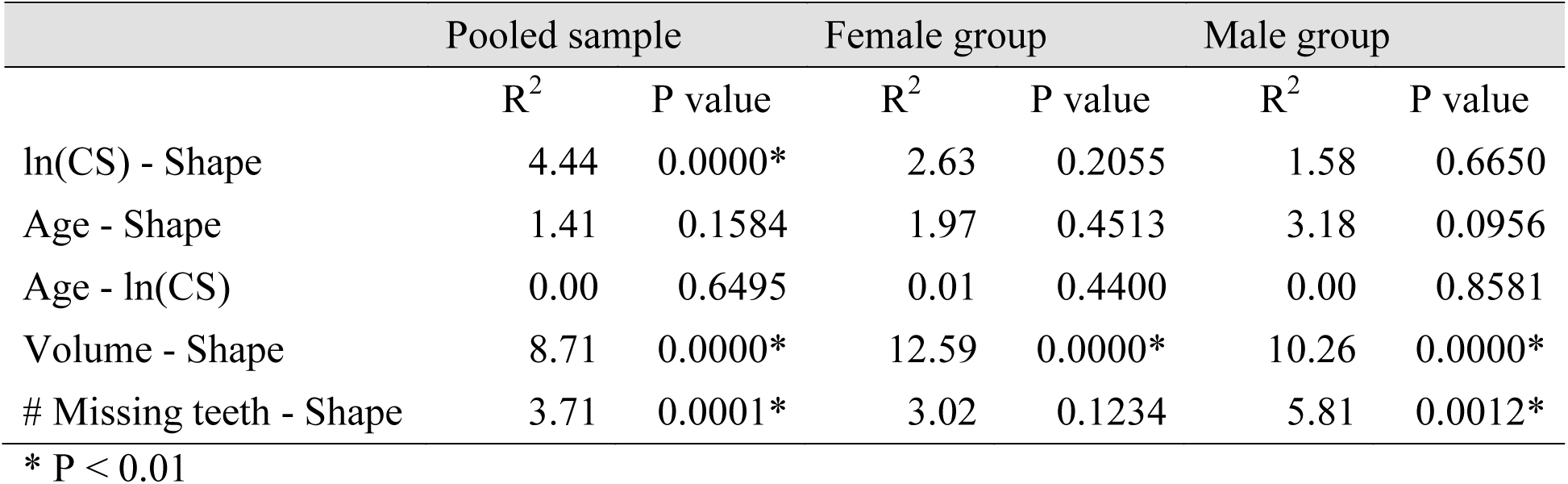
Correlation between variables. Correlations involving shape variables were tested with 10,000 permutations. Age - ln(CS) was computed using Pearson’s correlation coefficient.

|  | Pooled sample |  | Female group |  | Male group |  |
| --- | --- | --- | --- | --- | --- | --- |
|  | R<sup>2</sup> | P value | R<sup>2</sup> | P value | R<sup>2</sup> | P value |
| ln(CS) - Shape | 4.44 | 0.0000* | 2.63 | 0.2055 | 1.58 | 0.6650 |
| Age - Shape | 1.41 | 0.1584 | 1.97 | 0.4513 | 3.18 | 0.0956 |
| Age - ln(CS) | 0.00 | 0.6495 | 0.01 | 0.4400 | 0.00 | 0.8581 |
| Volume - Shape | 8.71 | 0.0000* | 12.59 | 0.0000* | 10.26 | 0.0000* |
| # Missing teeth - Shape | 3.71 | 0.0001* | 3.02 | 0.1234 | 5.81 | 0.0012* |

\* P < 0.01

### Age

There was no statistically significant correlation between age and shape, or between age and centroid size (Table 3).

### Ramus analysis

As expected, a clear size and shape dimorphism was also observed for the isolated ramus. Centroid size of males was larger by 11.8% (t-test, P < 0.0001). Shape differed significantly (P = 0.0003, 10,000 permutations). Similar shape differences were noted to those observed for the whole mandible (Figure 7). We could not observe gonial angle eversion in the male group, nor ramus flexure.

**Figure 7.**
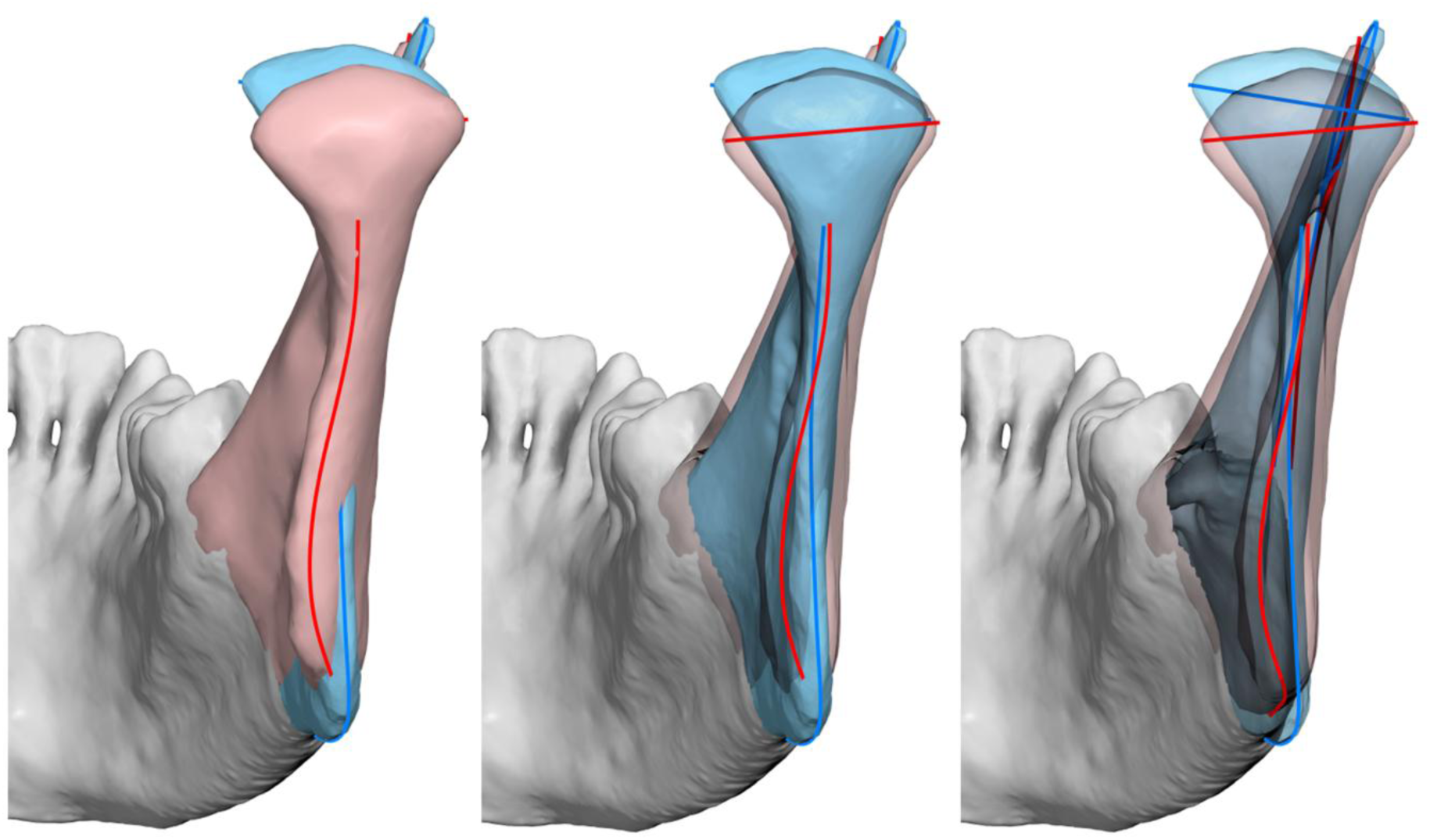
Superimposition of the male (blue) and female (red) groups showing shape dimorphism of the ramus (here exaggerated). Middle: female ramus transparent; right: both rami transparent. Note female outline (red) curving towards the lingual side, and difference in condylar axis angulation.

## Discussion

In creating an atlas of shape variability, the design of the template is a key factor [29]. The base mesh does not need to be detailed or be one of the meshes of the sample; indeed, simpler geometries sometimes work better [29]. We used a simplified symmetric mandibular mesh, expanded outwards by 1.5 mm, to avoid the common problem of the landmarks projecting on the wrong surface [29], especially in the area of the gonial angle, where the ramus can be thin and the gonial angle everted. We aimed for a large number of landmarks, dispersed evenly over the whole surface, to capture both the shape of the main ridges and the smooth surfaces in-between. The number of fixed landmarks was small, limited to the condyle poles, the coronoid processes, Gonion and Gnathion. Gonion is a problematic landmark, showing high identification error, both in 2D and 3D digitizations [33, 34, 35]. Although we could set this as a sliding semilandmark, or remove it altogether, we opted to retain it as a sentinel between the ramus and the corpus, to avoid the curve and surface semilandmarks from invading the wrong area. However, we located it algorithmically using a clearly defined geometric procedure (Supplementary Table 1), to reduce identification error. Gnathion was similarly located, as the farthest point from the condyle on the midsagittal plane. Locating landmarks algorithmically, based on their definitions, avoids subjectivity, and ensures repeatability and validity, although biological homology is debatable [36]. The curve semilandmarks were few and sparsely dispersed, to allow them to adjust by sliding, since a very high density effectively prohibits sliding and reverts to equidistant sampling. The surface semilandmarks were dispersed on the template mesh via a diffusion algorithm, to ensure an initially even distribution.

Alternatives to GM methods for constructing shape models are statistical shape models that establish correspondences of meshes at the vertex level [2, 3, 9]. Although highly dense models are produced, the anatomical correspondence (homology) is not guaranteed [13, 36]. We consider GM methods preferable, due to their solid statistical foundation and excellent visualization tools.

Our sample was a convenience sample, imaged for various reasons, most commonly for dental implant planning and third molar pre-extraction evaluation. Although there is no assurance that the average coincides with the average of the population, research has shown minimal effect of including even extreme cases [37]. Sample size was adequate for assessing average shape and shape variability [24, 25].

Landmarking was accurate, as shown by the repeated digitizations, because the curves were placed on well-defined ridges and the remaining surface points were located by a TPS warping of the template followed by sliding. Fixed landmarks were placed by automated heuristics (e.g. Gnathion, Gonion) further minimizing subjective identification [38]. The only curves that required full human intervention were the alveolar buccal and lingual curves. Almost 80% of the shape variance was described by the first 12 PCs, the first 3 of these describing 49%, so compactness was good and comparable to previous work [6, 16]. Van der Wel et al. [39] report a larger spread of shape variance among the PCs; this can be attributed to their sample comprising patients under treatment by orthognathic surgery, therefore potentially more extreme cases, and to segmentation artifacts in the area of the teeth, due to metallic orthodontic appliances. The number of landmarks is a significant factor that affects the percentage of shape variance that is collected and distributed in the principal components of shape. Studies with a few landmarks report a large fraction of variance in the first few PCs because shape is more coarsely measured (e.g. 14 landmarks: 67% shape variance in the first 2 PCs [40], 13 landmarks: 61% variance in the first 2 PCs [18, 19]).

The shape patterns were similar to those reported elsewhere, mainly related to mandibular width, angulation between the ramus and corpus, inclination of the symphysis and prominence of the gonial angle. PC1 described mandibular width variation, in relation to ramus height and corpus length (Figure 5) whereas PC2 mainly described the ramus-corpus angulation. The same primary patterns are seen in the work of van der Wel et al. [39], and potentially Fournier et al. [6] and Kim et al. [9], although the visualizations in those publications do not facilitate a direct comparison. The shape patterns obviously depend on several factors, including ethnicity, age, sex, and degree of edentulism. Our sample was mono-ethnic, equally divided by sex, and of low edentulism prevalence (average number of teeth missing: 2.2, Table 1), so the results need to be assessed under these conditions.

A significant effect of edentulism on mandibular shape has been observed [18, 19, 41], which we detected here, but only in the male group and the pooled sample. In addition to reduction in the height of the alveolar process, loss of teeth was associated with retraction of the anterior alveolar area with relative prominence of menton, increase of the gonial angle and intercondylar distance, and posterior inclination of the ramus (Supplementary Figure 5). We mention these associations with great caution, even though they agree with previous reports overall [18, 19, 41], since our sample contained very few patients with many (> 5) missing teeth, did not contain completely edentulous mandibles, and alveolar resorption had not progressed significantly in several patients.

A clear sexual dimorphism was evident, both regarding size and shape, as expected for an adult sample [42]. Centroid size, computed from the mesh vertices, was 8.9% larger in males; however, mesh volume was 25% larger. A discrepancy between the two is expected because, with scaling, volume increases to the third power, whereas centroid size to the first power.

However, the expected volume change would be larger, at 29% (1.089^3^ = 1.291). This can be explained by the mandible’s shape and the position of the centroid, which lies in empty space, on the midsagittal plane, at the level of the molars. The distance of the landmarks relative to the centroid is affected mostly by variation in mandibular width, and not so much by variation in ramus height, ramus anteroposterior width, or ramus and corpus thickness, factors that significantly affect volume. The superimposition of the size-adjusted averages (Figure 6) shows differences in shape that explain this discrepancy between volume and centroid size ratios, e.g. a higher ramus and a more pronounced gonial and symphyseal area in the males. Size dimorphism has been noted in all previous studies of adult samples.

Franklin et al. [16] reported almost the same centroid size difference (7.8%) for their sample of 30 mandibles. Vallabh et al. [43] list various linear measurements, of which ramus height shows the largest relative difference between sexes (14%) whereas width measurements are comparable to our centroid size ratio (intercondylar width: 5.6% and intergonial width 8.7%). This difference in intercondylar and intergonial widths is also reflected in the shape difference we detected (Figure 6). Kranioti et al. [44], in a sample of the same ethnic origin as ours, report a comparable inter-gonial width difference of 7.6%, giving a sex classification accuracy of 71%.

Shape dimorphism was less pronounced than size dimorphism, as seen when comparing Figure 4 and Supplementary Figure 4. In addition to a higher ramus, more pronounced gonial and mental areas, males showed a wider inter-gonial distance. Such differences between sexes have been noted by other investigators as well [16, 40].

One of the traits considered a male characteristic is gonial eversion, presumably arising from a strong masseteric attachment. However, evidence suggests that the dimorphism is lower than initially assumed [45, 46]. To overcome the qualitative nature of this trait, Oettlé et al. [46] applied GM methods confined to the posterior ramal and gonial areas and obtained quantitative data. Although they detected differences between males and females, mainly in the extent and location of the eversion, the accuracy of sexing was below 75%. Our sample showed a larger inter-gonial width in males (Figure 6), but there was no clear gonial eversion when examining the gonial area. On the contrary, the female outline curved towards the lingual and the male outline was straight (Figure 7). A difference in condylar axis angulation was also observed.

Another alleged dimorphic trait is ramus flexure, an “angulation of the posterior border of the mandibular ramus at the level of the occlusal surface of the molars” [47]. Although the initial results for this trait were very optimistic, later evidence has been more reserved [42, 45, 48, 49]. Unfortunately, this trait is qualitative and could not be incorporated into the GM analysis. Visual inspection of the posterior ramus border did not show flexure in our sample (Figure 7).

Discriminant analysis based on the first 2 or 4 PCs in form space was very successful in assigning subjects to their correct group (91% and 93%, respectively). However, this was achieved mainly due to size dimorphism, similarly to previous research using linear measurements between anatomical landmarks [42, 44]. Classification accuracy was higher than that reported by previous studies using conventional linear and angular measurements. Kranioti et al. [44] report an accuracy of 80% based on 2 linear variables in a Greek population. Other reports vary, from around 75% to almost 90%, using univariate or multivariate models [50, 51, 52, 53, 54, 55, 56, 57]. Franklin et al. [58] report an exceptionally high accuracy of 95%, but this needs to be interpreted with caution, as 10 variables were applied on a sample of 40 mandibles, suggesting a danger of overfitting. We did not detect an age-related shape change, in contrast to some previous reports, e.g., Costa-Mendes et al. [40]. As noted above, our sample size was relatively small in the higher and lower age bins; however, we had a similar degree of edentulism in both sex groups, whereas this was not recorded in [40] and could have biased the results.

### Limitations

The template has very few points on the condylar head, so variability in condylar form could not be investigated. This was outside the scope of the present investigation and requires GM analysis focused on that region.

The sample was of Greek ethnicity, therefore the results may not be generalizable to other ethnic groups.

Although age covered a wide range, there were few patients in the tail ends of the distribution.

Time of tooth loss was not available. The effect of tooth loss on alveolar bone resorption, and subsequent functional issues that depend on prosthetic rehabilitation, are expected to affect mandibular shape, but we could not evaluate these factors.

## Author contributions

D.H. designed the study, supervised the project and wrote the software for digitizing the images and analyzing the results. A.C. digitized the data, ran the analyses and prepared the visualizations. D.H. and A.C. interpreted and discussed the findings, and prepared the draft. D.H. and W.L. co-administered the ‘MARGO’ project and funding. All authors discussed the results and edited the final version of the paper. All authors have read and agreed to the published version of the manuscript.

## Data availability

The mandibular meshes used in this study are available from the Zenodo repository (https://doi.org/10.5281/zenodo.6335430). The template is available by author request.

## Competing interests

D.J.H. owns stock in dHAL Software, the company that markets Viewbox 4. Authors A.C. and W.L. declare no competing interests.

Correspondence and requests for materials should be addressed to D.J.H.

## Supplementary material

**Supplementary Table 1.** List of landmarks and curves. R: right, L: left. Total number of fixed landmarks: 9. Total number of semilandmarks: 496. Also see Figure 2.

| Curves |  |
| --- | --- |
| Notch (R & L) | From the tip of the coronoid to the anterior surface of the condyle. |
| Coronoid (R & L) | From the tip of the coronoid, along the anterior border of the ramus, to the level of the dentition. |
| Posterior inferior | From the posterior surface of the right condyle, along the posterior border of the ramus, past Gonion, along the inferior border of the mandible, to the symphysis, and similarly on the left side. |
| Symphysis | From the labial alveolar crest (infradentale), inferiorly around the symphysis, and up to the lingual alveolar crest (linguale). |
| Alveolar buccal | Along the alveolar buccal crest, from the midpoint of the second molar of one side to the corresponding point of the other side. In case of a missing molar, the position was estimated from the molar of the opposite side, or was estimated from the position of the remaining teeth. |
| Alveolar lingual | Similar to Alveolar buccal, but on the lingual side. |
| Landmarks (n = 9) |  |
| Condyle lateral (R & L) | The lateral pole of the condyle. |
| Condyle medial (R & L) | The medial pole of the condyle. |
| Coronoid (R & L) | The tip of the coronoid process. |
| Gonion (R & L) | The most posterior and inferior point at the gonial angle. Located as the farthest point of the gonial angle from the line connecting the condyle and Gnathion. |
| Gnathion | The most inferior and anterior point of the symphysis, on the midsagittal plane. Located as the farthest point from the condyle. |
| Semilandmarks sliding on curves (n = 84) |  |
| Notch | 5 semilandmarks on each of the Notch curves |
| Coronoid | 5 semilandmarks on each of the Coronoid curves |
| Posterior inferior | 31 semilandmarks on the Posterior inferior curve |
| Alveolar buccal | 12 semilandmarks on the Alveolar buccal curve |
| Alveolar lingual | 10 semilandmarks on the Alveolar lingual curve |
| Symphysis | 11 semilandmarks on the Symphysis curve |
| Semilandmarks sliding on the mesh surface (n = 426) |  |
| Exterior surface | 100 semilandmarks on the right side and 100 semilandmarks on the left side, on the buccal/labial surface of the mandible. |
| Interior surface | 80 semilandmarks on the right side and 80 semilandmarks on the left side, on the lingual surface of the mandible. |
| Condyle Coronoid medial | 26 semilandmarks on the right side and 26 semilandmarks on the left side, on the medial surface of the condylar and coronoid processes. |
| Condylar head | 7 semilandmarks on each condylar head. |

**Supplementary Table 2.**
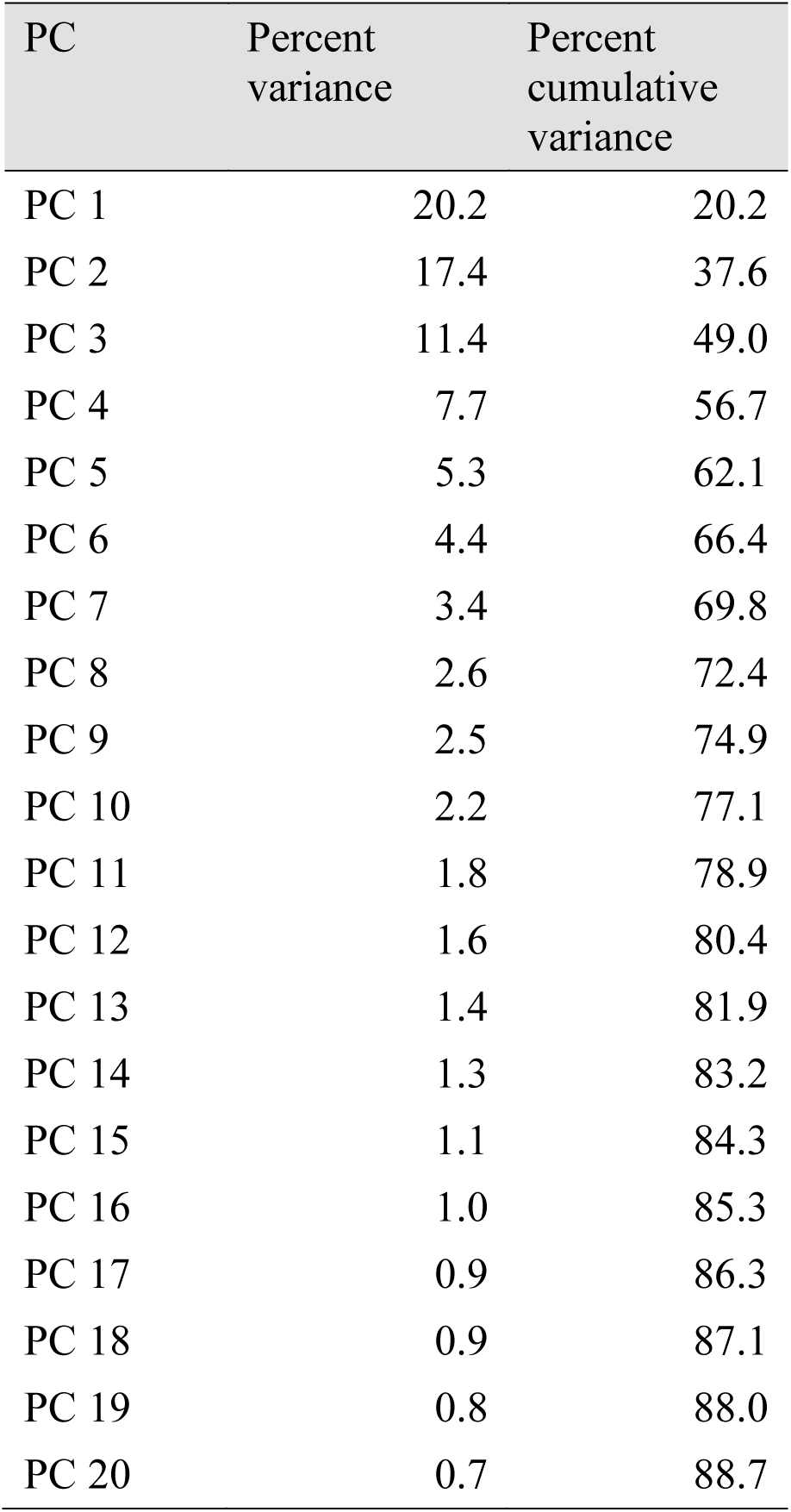
Percent variance described by the first 20 principal components in shape space for the whole sample.

| PC | Percent variance | Percent cumulative variance |
| --- | --- | --- |
| PC 1 | 20.2 | 20.2 |
| PC 2 | 17.4 | 37.6 |
| PC 3 | 11.4 | 49.0 |
| PC 4 | 7.7 | 56.7 |
| PC 5 | 5.3 | 62.1 |
| PC 6 | 4.4 | 66.4 |
| PC 7 | 3.4 | 69.8 |
| PC 8 | 2.6 | 72.4 |
| PC 9 | 2.5 | 74.9 |
| PC 10 | 2.2 | 77.1 |
| PC 11 | 1.8 | 78.9 |
| PC 12 | 1.6 | 80.4 |
| PC 13 | 1.4 | 81.9 |
| PC 14 | 1.3 | 83.2 |
| PC 15 | 1.1 | 84.3 |
| PC 16 | 1.0 | 85.3 |
| PC 17 | 0.9 | 86.3 |
| PC 18 | 0.9 | 87.1 |
| PC 19 | 0.8 | 88.0 |
| PC 20 | 0.7 | 88.7 |

**Supplementary Figure 1.**
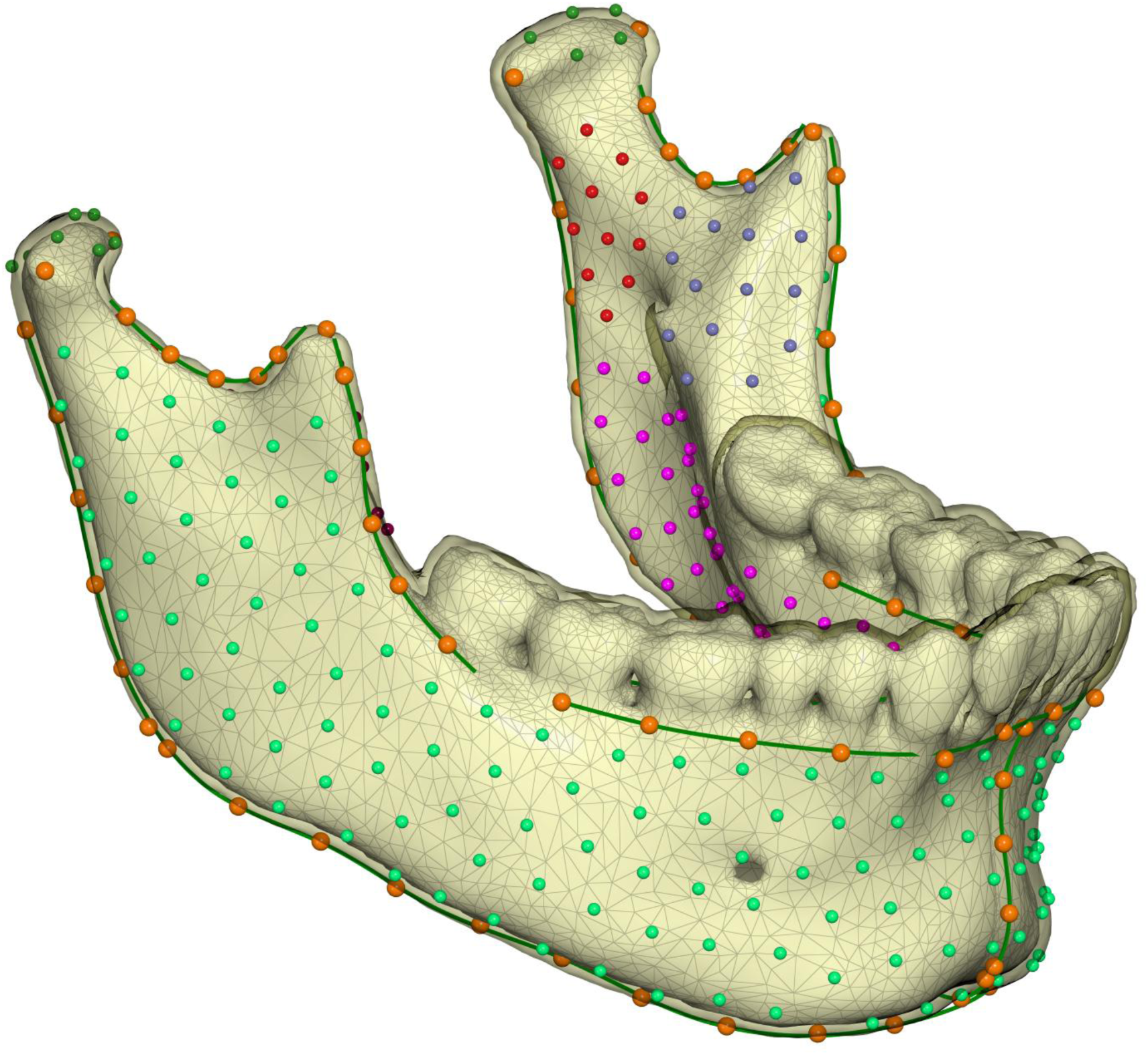
The template of curves and landmarks, overlaid on a symmetric simplified mesh, expanded by 1.5 mm (transparent overlay) to ensure correct landmark projection when transferring points, mainly those on the medial and lateral ramus surfaces.

**Supplementary Figure 2.**
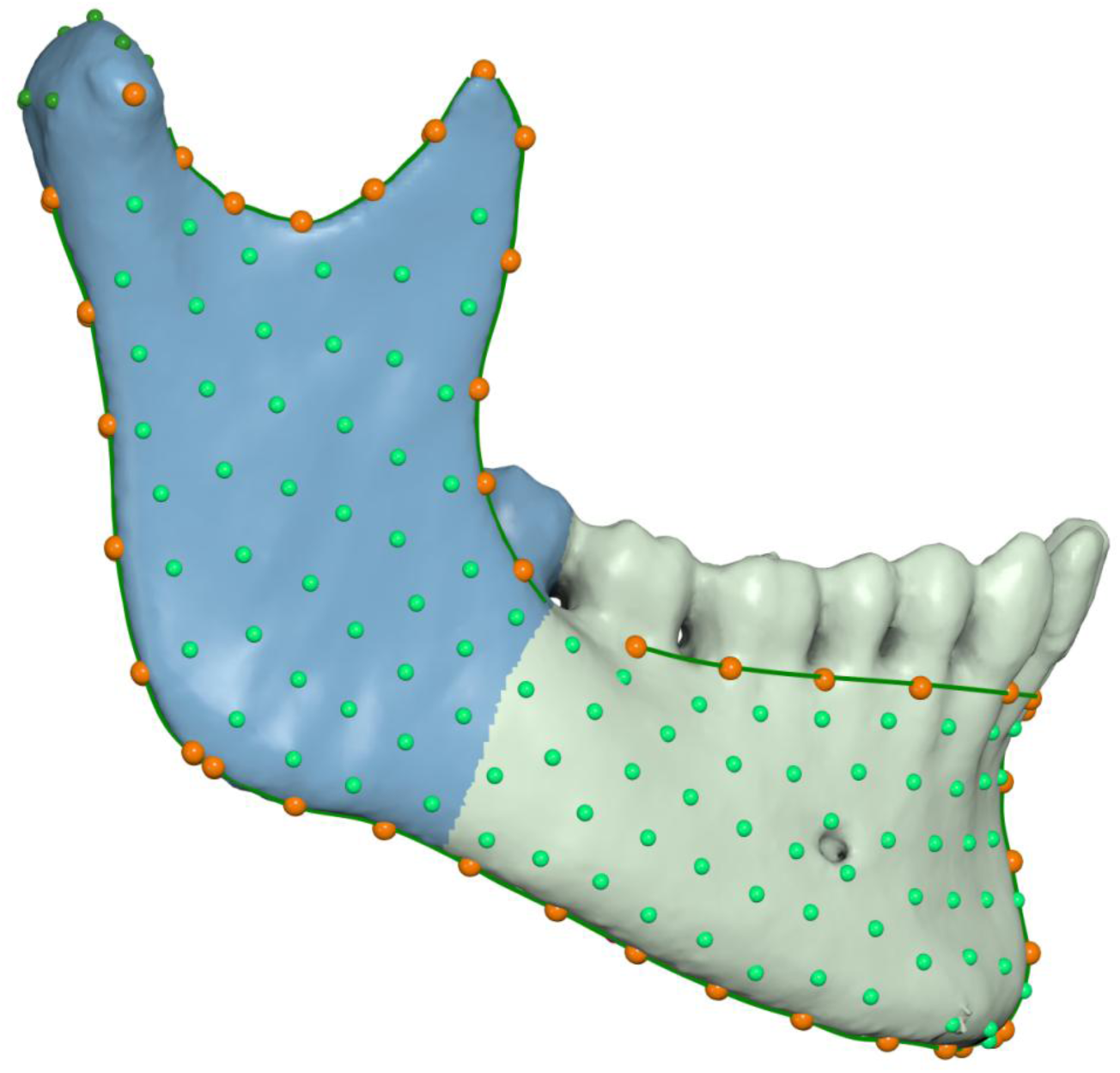
Landmarks included for the ramus analysis are shown in the blue region.

**Supplementary Figure 3.**
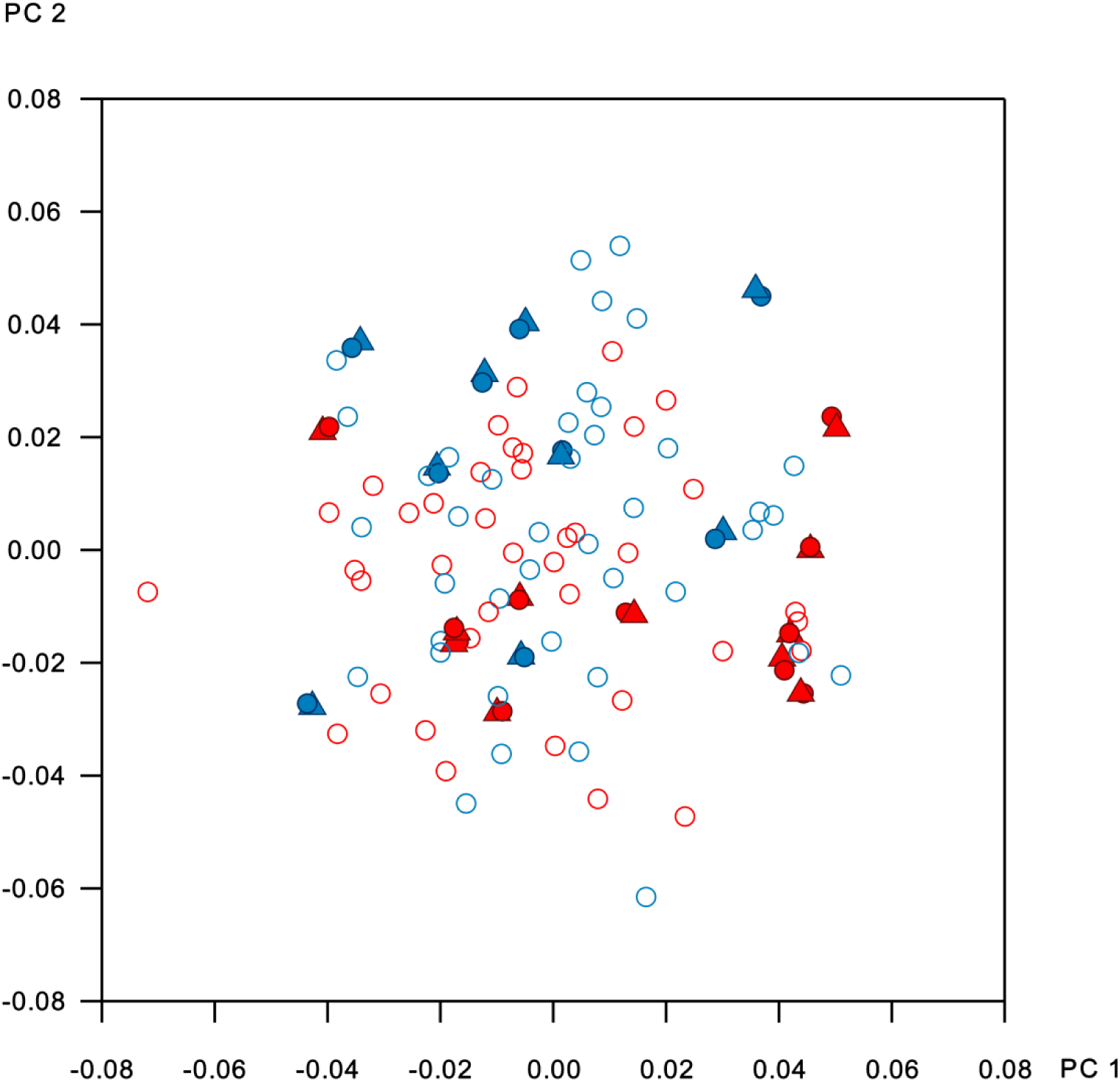
Repeated digitizations (filled triangles) in shape space, showing small error relative to the original digitizations (filled circles) and overall sample variability. Open circles: non-repeats. Blue: male, red: female.

**Supplementary Figure 4.**
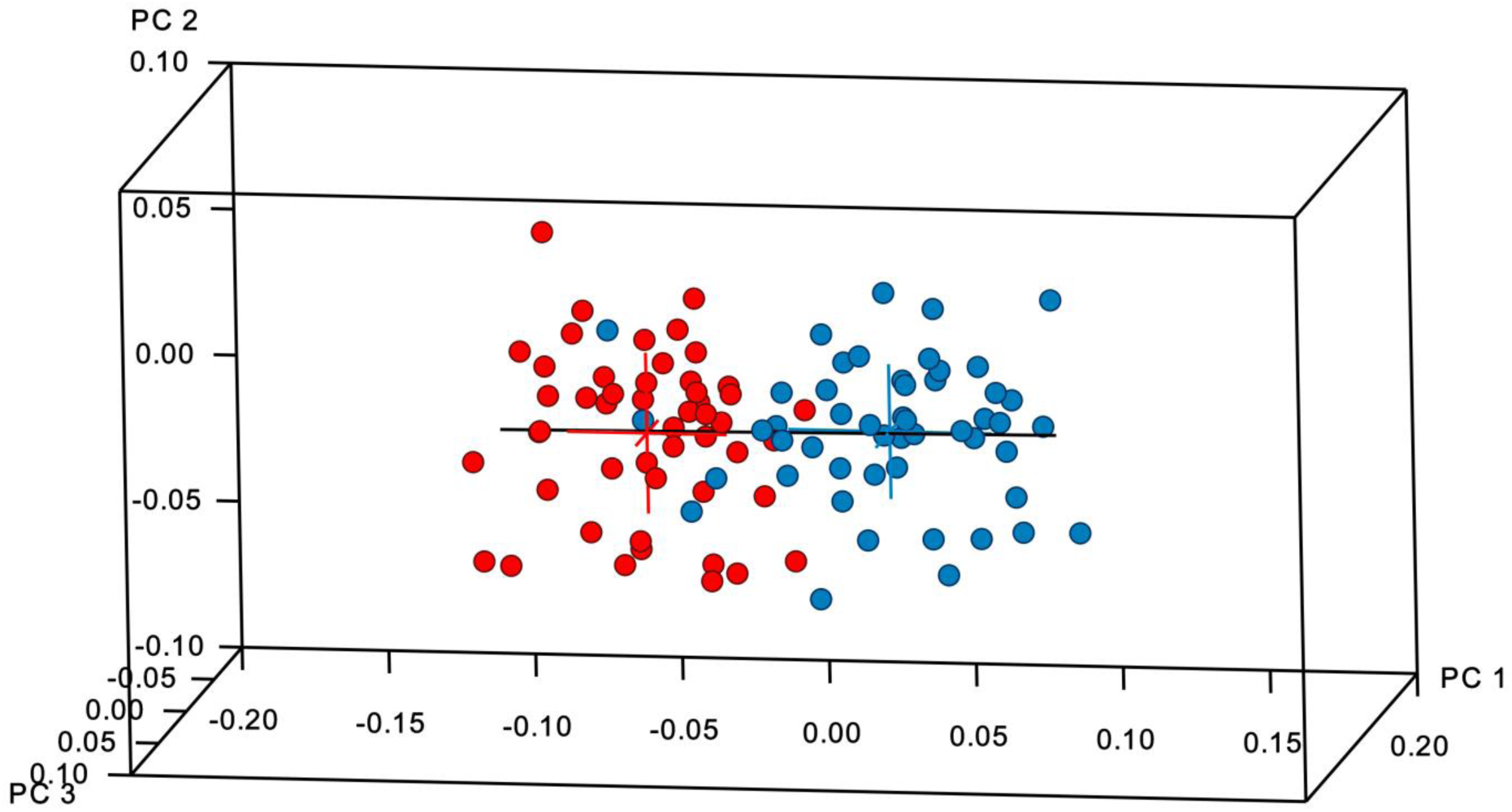
Plot of the sample in form space. Blue: males, red: females. Black line: vector of pure size variance.

**Supplementary Figure 5.**
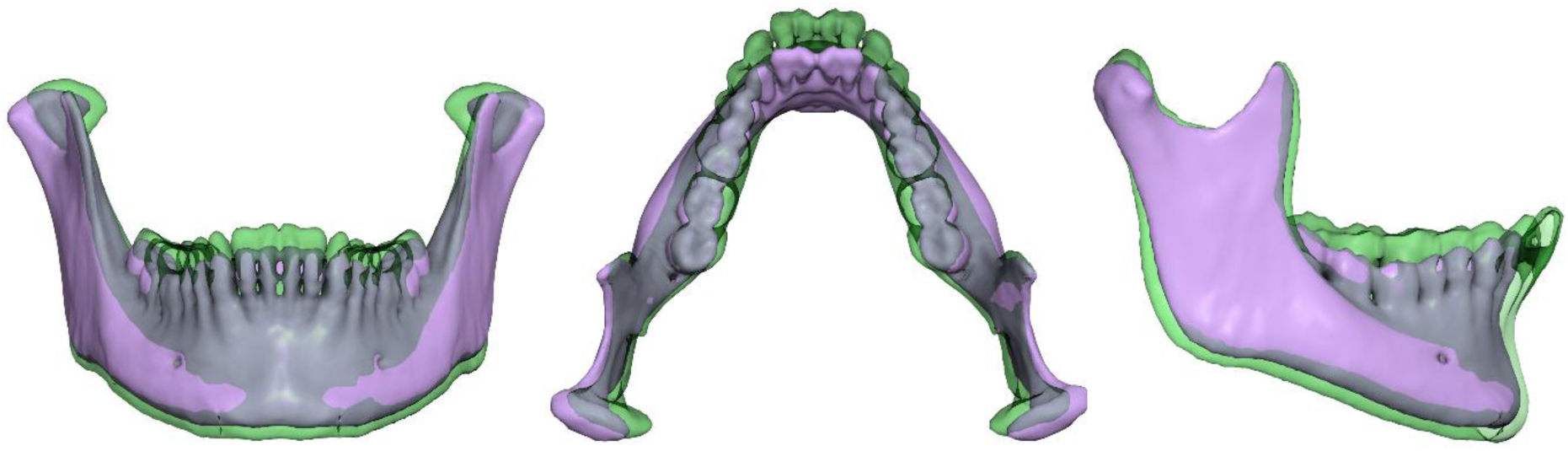
Superimposition of mandibular shape warped to ±3 standard deviations along the regression vector of shape on number of missing teeth for the pooled sample. Green: dentate; purple: edentulous. Here, the same exemplar shape has been warped, so teeth are visible in both extremes.

